# Applying Benford’s law to COVID-19 data: The case of the European Union

**DOI:** 10.1101/2021.12.24.21268373

**Authors:** Pavlos Kolias

## Abstract

Previous studies have used Benford’s distribution to assess whether there is misreporting of COVID-19 cases and deaths. Data inaccuracies provide false information to the media, undermine global response, and hinder the preventive measures taken by countries worldwide. In this study, daily new cases and deaths from all the countries of the European Union were analyzed and the conformance to Benford’s distribution was estimated. For each country, two statistical tests and two measures of deviation were calculated to determine whether the reported statistics comply with the expected distribution. Four country-level developmental indexes were also included, the GDP per capita, health expenditures, the Universal Health Coverage index, and the full vaccination rate. Regression analysis was implemented to show whether the deviation from Benford’s distribution is affected by the aforementioned indexes. The findings indicate that four countries were in line with the expected distribution, Bulgaria, Croatia, Lithuania, and Romania. For the daily cases, Denmark, Greece, and Ireland, showed the greatest deviation from Benford’s distribution and for deaths, Malta, Cyprus, Greece, Italy, and Luxemburg exhibited the highest deviation from Benford’s law. Furthermore, it was found that the vaccination rate is positively associated with deviation from Benford’s distribution. These results suggest that overall, official data provided by authorities are not confirming Benford’s law, yet this approach is not conclusive; it acts as a preliminary tool for data verification. More extensive studies should be made with a more thorough investigation of countries that showed the greatest deviation.

## Introduction

The pandemic of COVID-19 has affected the life of millions of people worldwide. Due to the rapid contagiousness of the virus (Hafeez et al., 2020), nearly every country employed measures against the virus’ spread, such as national lockdowns and restrictions of outdoor activities. The pandemic showed that statistical and machine learning modelling can potentially predict the number of new cases or deaths for a given country (Cássaro & Pires, 2020; Niazkar & Niazkar, 2020; Neto et al., 2020). The accurate forecast of the infection curve can facilitate government’s measures towards the suppression of the growth rate. However, in order to accurately predict or model COVID-19 spread, reliable and valid data should be collected from authorities. The recent pandemic of COVID-19 raised issues about data collection and handling. Media reports have questioned whether the statistics provided by countries are trustworthy (Kilani, 2021). Several studies have disputed the accuracy of government data and had linked data manipulation with transparency and democracy indexes (Adsera, Boix & Payne, 2003; Magee & Doces, 2015; Rozenas & Stukal, 2019).

Previous studies, in different fields, have applied Benford’s distribution (or law) analysis to detect fraudulent and manipulated data. Specifically, for COVID-19, it was found that deaths were underreported in the USA (Campolieti, 2021), while in China no manipulation was found (Koch & Okamura, 2020). A study for Japan also showed deviation from Benford’s distribution (Lee, Han & Jeong, 2020). Furthermore, it was found that countries with higher values of the developmental indexes are less likely to deviate from Benford’s law (Balashov, Yan & Zhu, 2021).

This study applies Benford’s law to detect the first digit deviations of the announced cases and deaths from the expected frequencies in the European Union (EU). We further investigate whether the estimated deviation for each country, is associated with four developmental indexes, the GDP per capita, health expenditures (% of GDP), the Universal Health Coverage Index and full vaccination rate.

## Methods

### Sample

The public COVID-19 data of the European Union, regarding daily cases and deaths were exported from the European Centre for Disease Prevention and Control (ECDC) and consisted of observations between 2^nd^ of March to the 20^th^ of December 2021 (*N* = 8820). ECDC’s Epidemic Intelligence team collects and refines daily data of new cases and deaths associated with COVID-19, based on reports from health authorities worldwide. Apart from COVID-19 data, we included the gross domestic product per capita (GDPc), the healthcare expenditures of countries as percentage of GDP (HGDP), and the Universal Health Coverage Index (UHC) from the World Bank (https://data.worldbank.org/). Finally, we included the full COVID-19 vaccination rate as of the 16^th^ of December 2021, obtain from ECDC.

### Benford’s distribution

Benford’s law (or law of prime digits) is a probability distribution for determining the first digit in a set of numbers. It was formally proposed in 1938, after an early work by the mathematician Simon Newcomb, and by the physicist Frank Benford, who claimed that in natural and unrestricted data sets, the probability of each digit appearing is given by the formula:

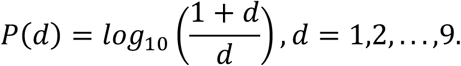

Based on Benford’s distribution, the probabilities for each number *d* as the first digit are presented in Table 1.

**Table 1.** Probabilities of the digit in the first position

| Digit | 1 | 2 | 3 | 4 | 5 | 6 | 7 | 8 | 9 |
| --- | --- | --- | --- | --- | --- | --- | --- | --- | --- |
| $p$ | .301 | .176 | .125 | .097 | .079 | .067 | .058 | .051 | .046 |

Note: The probabilities are rounded to 3 decimal places.

The most common application of the law is in Economics, where it has already been considered as a tool for checking tax validity and detecting fraud (Nigrini, 1996; Durtschi, Hillison & Pacini, 2004; Tam Cho & Gaines, 2007). More recent studies have used Benford’s law to investigate whether COVID-19 data provided by countries are accurate (Kilani, 2021; Silva & Figueiredo Filho, 2021; Campolieti, 2021; Koch & Okamura, 2020) and if the deviation from Benford’s distribution could be affected by developmental indexes (Balashov, Yan & Zhu, 2021). Conformity with Benford’s distribution is expected when the data under consideration spread over several terms of magnitude, however for countries with limited cases and/or deaths, the distribution of the first digit would generally deviate from Benford’s.

### Goodness-of-fit

First, in order to investigate to which extent, the observed cases and deaths conform to Benford’s law’s expected frequencies, two goodness-of-fit tests were applied, the chi-squared (χ^2^) goodness-of-fit test and Kolmogorov-Smirnov (K-S). The chi-squared test statistic is given by:

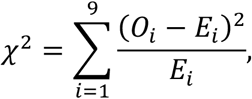

where the index *i* is the digit, and *O*_*i*_ and *E*_*i*_ are the observed and expected frequencies of the i-th digit, respectively. The degrees of freedom for this test are equal to 8, and the critical value is 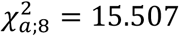 for the significance level set at *a* = 0.05; thus, any value of the statistic greater than the critical value would imply significant deviation from the expected distribution. However, in large samples, the interpretation of significance should be avoided, as the test has enough power to detect even small deviations from the expected distribution (Lin, Lucas & Shmueli, 2013). To accompany the results of the chi-squared test, Cramer’s V was calculated along with 95% bootstrap confidence interval (CI) for an estimate of the effect size. The Kolmogorov-Smirnov *D* statistic is commonly used for comparing empirical with theoretical continuous distributions, but it can also be used with integers. The statistic is given by:

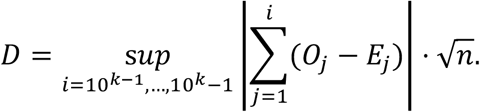

Both chi-squared and D statistics are greatly affected by sample size, hereby we included two measures that are not affected by large sample sizes, namely the Euclidean distance (*ED*) in the nine-dimensional space (Tam Cho & Gaines, 2007) given by:

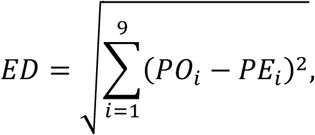

and Mean Absolute Distance (MAD) given by:

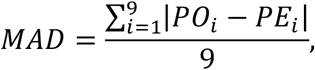

where *PO*_*i*_ and *PE*_*i*_ are the observed and expected proportions of the first digit, respectively. As 27 countries of the EU were tested for conformity with Benford’s law, familywise error rate was also considered and the Holm-Bonferroni correction was applied to adjust the p-values obtained (Aickin & Gensler, 1996).

### Regression analysis

The two measures of deviation (ED and MAD) were used as the dependent variables in two regression models, with independent variables the gross domestic product per capita (GDPc), the healthcare expenditures of countries as percentage of GDP (HGDP), the Universal Health Coverage Index (UHC) and the full COVID-19 vaccination rate (Vac), to examine whether the distance observed from Benford’s distribution could be associated with those predictors. Instead of relying in ordinary least squares (OLS) estimates for the parameters of the model, bootstrap estimates have been calculated due to the small sample size of countries (N = 27) leading to more robust results. With bootstrap, 10000 samples were selected, and each time we estimated the OLS coefficients of the parameters; hence, creating the sampling distribution of each coefficient along with 95% bootstrap CIs (Davison & Hinkley, 1997).

## Results

The results of the goodness-of-fit tests along with the two measures of deviations are presented in Table 2. For almost all countries, except for Bulgaria, Croatia, Lithuania and Romania, significant deviations from Benford’s distribution were found for both cases and deaths. For daily cases, Denmark, Ireland, and Greece were associated with the highest chi-squared statistics and this was also confirmed by the two distance measures (Figure 1 and 2). Regarding deaths, Cyprus, Italy, and Greece had the highest chi-squared statistics and distance measures. The K-S D statistic in most cases came in agreement with the chi-squared test.

**Table 2.** Goodness-of-fit statistics and distance measures across countries for new cases and deaths associated with COVID-19

|  | Cases |  |  |  | Deaths |  |  |  |
| --- | --- | --- | --- | --- | --- | --- | --- | --- |
| | $\chi^2$ | D | ED | MAD | $\chi^2$ | D | ED | MAD |
| Austria | 30.49** | 2.45*** | 1.905 | 0.032 | 13.29 | 1.57* | 1.361 | 0.018 |
| Belgium | 24.19* | 1.33 | 1.846 | 0.022 | 45.99*** | 1.90** | 2.53 | 0.038 |
| Bulgaria | 12.00 | 0.90 | 1.072 | 0.016 | 12.43 | 0.90 | 1.262 | 0.020 |
| Croatia | 8.00 | 0.90 | 1.202 | 0.018 | 13.46 | 0.85 | 1.148 | 0.020 |
| Cyprus | 7.31 | 1.30 | 1.03 | 0.016 | 75.12*** | 6.36*** | 3.051 | 0.060 |
| Czech Republic | 16.87 | 1.73* | 1.746 | 0.024 | 17.14 | 3.03*** | 1.643 | 0.023 |
| Denmark | 110.34*** | 4.37*** | 3.589 | 0.057 | 40.05*** | 4.61*** | 2.034 | 0.040 |
| Estonia | 15.62 | 1.01 | 1.3 | 0.021 | 20.31 | 6.30*** | 1.681 | 0.033 |
| Finland | 25.56* | 3.15*** | 1.74 | 0.031 | 30.25** | 7.23*** | 1.625 | 0.038 |
| France | 36.51*** | 1.49 | 2.32 | 0.036 | 7.71 | 0.67 | 1.041 | 0.017 |
| Germany | 5.62 | 0.61 | 0.917 | 0.015 | 24.29* | 1.14 | 2.005 | 0.029 |
| Greece | 77.20*** | 2.33*** | 3.389 | 0.050 | 54.84*** | 3.52*** | 3.108 | 0.045 |
| Hungary | 15.71 | 3.43*** | 1.13 | 0.018 | 33.79** | 4.08*** | 2.273 | 0.041 |
| Ireland | 99.48*** | 2.58*** | 3.075 | 0.054 | 10.08 | 11.90*** | 0.798 | 0.025 |
| Italy | 10.08 | 0.44 | 1.11 | 0.018 | 60.58*** | 2.71*** | 3.303 | 0.045 |
| Latvia | 31.05** | 1.59 | 1.732 | 0.027 | 5.89 | 2.51*** | 1.081 | 0.015 |
| Lithuania | 4.62 | 0.38 | 0.694 | 0.011 | 22.68 | 1.44 | 1.886 | 0.028 |
| Luxembourg | 21.66 | 4.35*** | 1.876 | 0.030 | 30.77** | 10.96*** | 2.347 | 0.055 |
| Malta | 15.29 | 1.31 | 1.25 | 0.020 | 50.94*** | 11.90*** | 2.966 | 0.079 |
| Netherlands | 61.24*** | 1.37 | 2.926 | 0.047 | 12.49 | 0.47 | 0.925 | 0.016 |
| Poland | 29.73** | 2.03*** | 1.932 | 0.032 | 25.43* | 2.04** | 1.755 | 0.027 |
| Portugal | 17.74 | 1.78** | 1.952 | 0.024 | 25.89* | 1.79** | 1.811 | 0.030 |
| Romania | 9.61 | 0.76 | 1.036 | 0.014 | 11.17 | 0.90 | 1.32 | 0.018 |
| Slovakia | 9.22 | 1.02 | 1.003 | 0.016 | 28.22** | 4.42*** | 1.66 | 0.029 |
| Slovenia | 17.37 | 1.02 | 1.154 | 0.019 | 12.47 | 3.65*** | 1.524 | 0.021 |
| Spain | 18.94 | 5.25*** | 1.492 | 0.028 | 7.16 | 5.25*** | 1.256 | 0.018 |
| Sweden | 17.87 | 1.95* | 1.219 | 0.022 | 19.32 | 2.04** | 1.525 | 0.025 |

\*\*\* $p < .001$ , \*\* $p < .01$ , \* $p < .05$ , $p$ -values were adjusted with the Holm-Bonferroni method.

**Figure 1.**
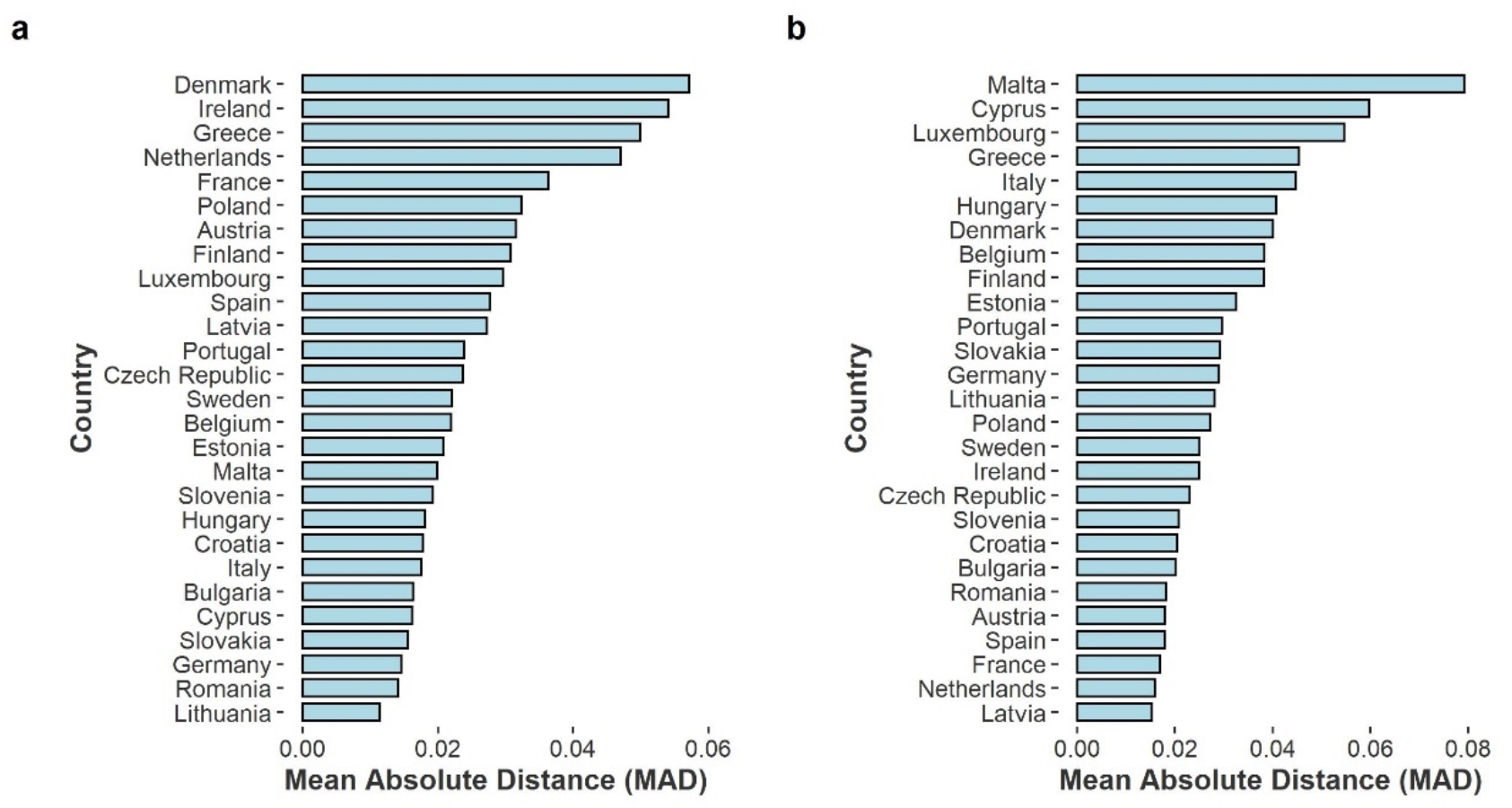
Mean Absolute Distance across countries for a) cases and b) deaths.

**Figure 2.**
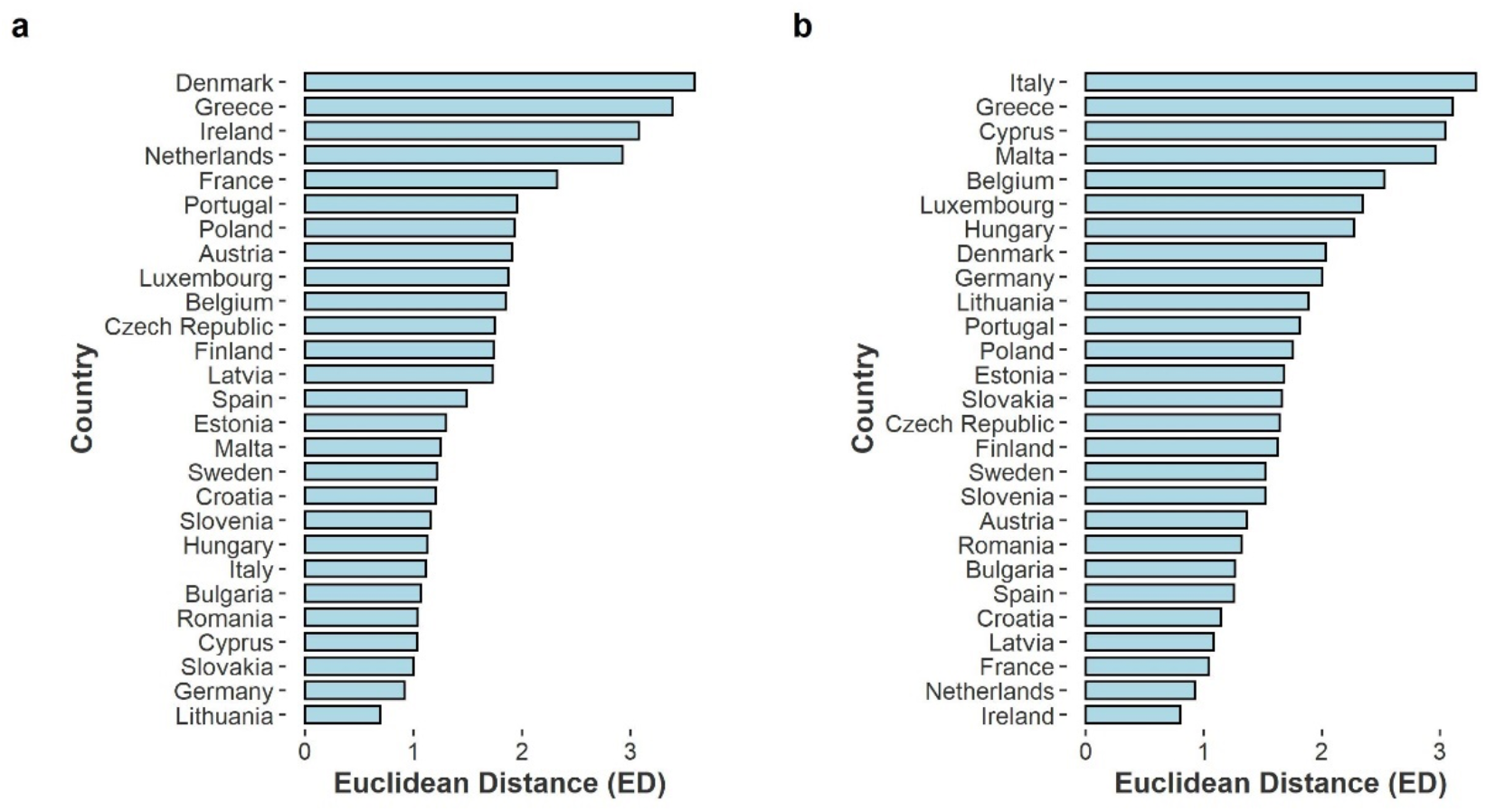
Euclidean Distance across countries for a) cases and b) deaths.

The bootstrap estimates and 95% bootstrap CIs of the regression analysis for the two measures of deviation are presented in Table 3. In order to avoid having small coefficients, GDP per capita has been log-transformed and the other three predictors were divided by 100. Regarding new cases, no predictor was found to significantly affect either MAD or ED. Vaccination rate was positively associated with deviation from Benford’s distribution in deaths as measured by MAD (0.076, 95% CI [0.020, 0.144]) and as measured by ED (3.415, 95% CI [1.175, 6.286]), indicating that countries with a higher full vaccination percentage tend to deviate more from Benford’s law.

**Table 3.**
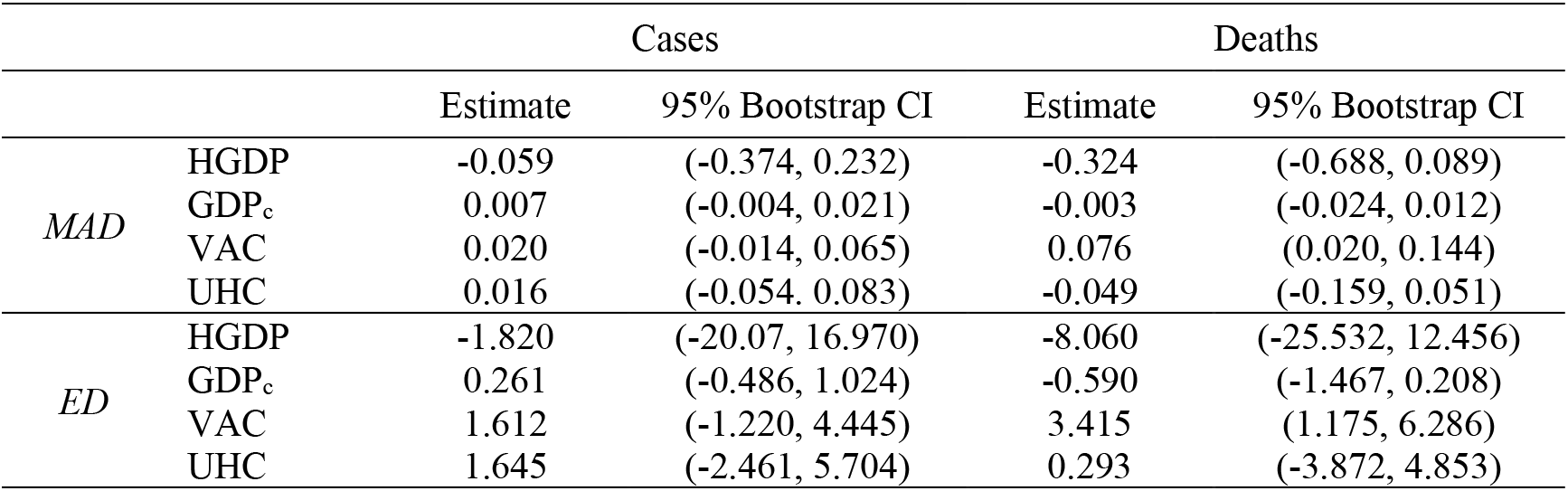
Bootstrap estimates and 95% CIs for Mean Absolute Deviation and Euclidean Distance from Benford’s distribution

## Discussion

This study aimed to apply Benford’s law to COVID-19 data from the European Union. Data of daily new cases and deaths were collected by ECDC for the period of the 2^nd^ of March 2021 and 20^th^ of December 2021. Also, four country-level indexes were collected, the GDP per capita, the health expenditure as GDP percentage, the Universal Health Coverage index and the full vaccination rate. Two goodness-of-fit tests were applied, the chi-squared test and the Kolmogorov-Smirnov test, and two measures of deviation were estimated, the Euclidean distance and Mean Absolute distance. Bulgaria, Croatia, Lithuania, and Romania were not deviating from Benford’s law for both new cases and deaths. Regarding daily cases, Cyprus, Estonia, Germany, Italy, Malta, Slovakia, and Slovenia were in line with Benford’s distribution, while Denmark, Greece and Ireland, showed the greatest distance from Benford’s distribution. Regarding deaths, the chi-squared test showed that France, Ireland, Latvia, Netherlands, Slovenia and Spain matched Benford’s law whilst Malta, Cyprus, Greece, and Italy had the highest distance from Benford’s distribution. These results do not necessarily indicate fraudulent data, especially for deaths, as countries with a small number of daily new deaths are not expected to behave according to Benford’s law. The results from the regression analysis suggested that the full vaccination rate was positively associated with non-conformity with Benford’s law, where countries with the highest vaccination percentage exhibited greater deviation. This may imply that countries with a higher vaccination rate were also associated with fewer deaths, thus the data did not confirm Benford’s law.

The results of this study imply that the deviation from Benford’s law is not associated with country’s economy, which was suggested by earlier findings (Hollyer, Rosendorff & Vreeland, 2011). However, the effect would possibly be more apparent by including developing along with developed countries (Judge & Schechter, 2009). Deviations from Benford’s distribution do not imply that data are fraudulent, but this approach may act as a preliminary step for obtaining evidence for data manipulation; it is suggested that for specific economies that showed the greatest deviations, further studies could be made validating data reported by authorities. Additional parameters can be included, such as lockdown restrictions, preventive measures, and regional statistics and indicators.

## Data Availability

All data produced are available online at: https://data.worldbank.org/

